# Plasma protein profiling of multiple sclerosis using proximity extension assays

**DOI:** 10.1101/2022.07.29.22278211

**Authors:** Jesse Huang, Mohsen Khademi, Fredrik Piehl, Tomas Olsson, Ingrid Kockum

## Abstract

**Background:** Multiple sclerosis (MS) is an inflammatory disease characterized by demyelination and neuro-axonal degeneration in the central nervous system. Except for neurofilament light protein, identification of biomarkers has been difficult to assess in the blood, presumably due partly to sensitivity. To detect traces of disease activities in the periphery and identify low-abundance protein biomarkers, this study conducts an exploratory examination of the plasma proteome of MS using proximity extension technology, a high-sensitivity multiplex PCR-based immunoassay.

**Methods:** A case-control cohort consisting of 52 MS cases (relapsing-remitting=30, progressive=22) and 17 healthy controls were enrolled at the Karolinska University Hospital. EDTA plasma was analyzed for 1157 unique protein targets across thirteen proximity extension assays. Protein associations to disease outcomes and related clinical measures were assessed using a multivariable linear regression model corrected for sex and age at sampling.

**Results:** AHCY and CHR levels were higher among MS cases than controls, while FABP2 was lower among those with relapsing-remitting disease than controls (P_discovery_<0.05, P_replication_<0.05), although not significant after multiple test corrections. Furthermore, PTN and CYR61 levels were higher in progressive MS than in relapsing-remitting disease (P<0.0002, P_FDR_<0.05), and CRNN and CXCL13 were associated with more severe disability at sampling (P<0.0001, P_FDR_<0.05), independent of disease course. CTSF was positively correlated with disease duration (P=4.1×10^−5^, P_FDR_=0.044), while RRM2B level correlated with intrathecal immunoglobulin production (IgG Index) in relapsing-remitting MS (P=1.7×10^−5^, P_FDR_=0.018).

**Conclusion:** We provide several candidates for characterizing MS, particularly progressive disease, which may help monitor disease progression and treatment response in a clinical setting.

## Introduction

Multiple sclerosis (MS) is a complex disease typically featuring recurring inflammatory attacks in the central nervous system (CNS), leading to impaired neurological functions between periods of remission (relapsing-remitting). However, onset may begin as (primary) or transition into (secondary) a gradual worsening of neurological disability without remission (progressive). Although the exact cause of MS is unclear, it is well accepted that pathogenic lymphocyte activation in the periphery and infiltration through the blood-brain barrier cause localized inflammation and subsequent demyelination and neurodegeneration in the CNS^1^. Due to proximity to the target organ, the resulting inflammation and axonal damage might best be reflected in the cerebrospinal fluid (CSF)^2-4^. However, lumbar puncture is an invasive measure not suitable for monitoring disease activity and treatment response. Despite the many gene/protein candidates, it has been challenging to identify effective diagnostic and prognostic MS biomarkers in the blood^4-7^. However, recent technical advancements with access to high-sensitivity methodologies allow the detection of minute protein concentrations present in the periphery^8^.

This study utilizes proximity extension technology^9^, a high-throughput immunoassay based on oligonucleotide antibodies and qPCR amplification for high-sensitivity protein quantification. We have previously identified several MS-associated biomarkers using a small targeted panel of inflammation-related proteins with the same technology^4^. Here, we conducted a broader exploration of the plasma proteome (>1100 proteins) to identify additional peripheral biomarkers for MS and associated clinical characteristics.

## Methods

Blood was sampled from MS cases (n=52) with relapsing-remitting (RRMS, n=30), primary progressive (PPMS, n=7), or secondary progressive disease (SPMS, n=15) at the Karolinska University Hospital (Stockholm, Sweden)^2^. Disability and rate of progression were assessed using the expanded disability status scale (EDSS) and MS severity score (MSSS) by a qualified neurologist. Cases were compared to healthy controls (HC, n=17) with no known neurological disorders. The study was approved by the Stockholm Regional Ethical Review Board (no. 2009/2107-31/2, 2015/1280-32), and all participants have provided informed and written consent.

Relative protein concentration was measured using proximity extension assays (PEA) with thirteen *Olink Target 96-plex* panels for 1157 unique protein targets (**S-Table 1**). Protocols for the assay and data preprocessing have been detailed previously^4,9^. In summary, samples were incubated with a mixture of paired oligonucleotide antibody probes. The required matching of unique DNA reporter pairs for hybridization limits cross-reactivity before quantification with qPCR as log base-2 normalized protein expression (NPX) values. Internal assay controls assessed sample variability, and measures were then filtered by >60% call rate (i.e., the proportion above the detection limit).

Protein associations to MS disease and clinical course were assessed using a multivariable linear regression model adjusting for sex and age at sampling. Proteins with enriched tissue expression in the brain were first identified and examined as potential neuron/axonal damage markers (See **S-Table1**). Primary associations were initially identified by cross-validation using a discovery/replication partition (See **S-Figure 1**). Additional associations to common clinical measures such as EDSS (≥ 3), MSSS (≥ 5), IgG index (>0.77), CSF mononuclear cell count (>2×10^6^/L), the number of T2 MRI lesions (>8), and CSF neurofilament light chain (NEFL) [2] were examined among MS cases. Sensitivity analyses correcting for the sampling period and established handling markers (e.g., CD40L, AXIN1) were performed to check the reliability of primary findings^10^.

## Results

AHCY and CRH levels were higher among MS cases in both discovery and replication partitions, while FABP2 was lower among RRMS cases than healthy controls (P_discovery_<0.05, P_replication_<0.05) (**Figure 1, S-Figure 2**). MS cases also had suggestively higher LTA4H, SEMA4C, GLRX, IL15, and AOC1 and lower PARP1, CCL15, and DCTPP1 (P<0.01). However, associations were not significant after multiple test corrections in the overall cohort. Although not associated with MS, PTN and CYR61 levels were significantly higher among those with progressive rather than relapsing-remitting disease (P<0.0002, P_FDR_<0.05). Additional associations to progressive disease were observed with IDUA, GUSB, FOLR1, and CA3. Several proteins with enriched tissue expression in the brain were associated with MS, including CRH and MDGA1 (β=0.58, P=0.038). MOG levels were lower among secondary progressive than relapsing-remitting disease (β=-0.33, P=0.034). MS cases also had lower MEPE and BCAN levels and higher NEFL levels; however, these associations were not significant after correcting for sex and age at sampling.

**Figure 1.**
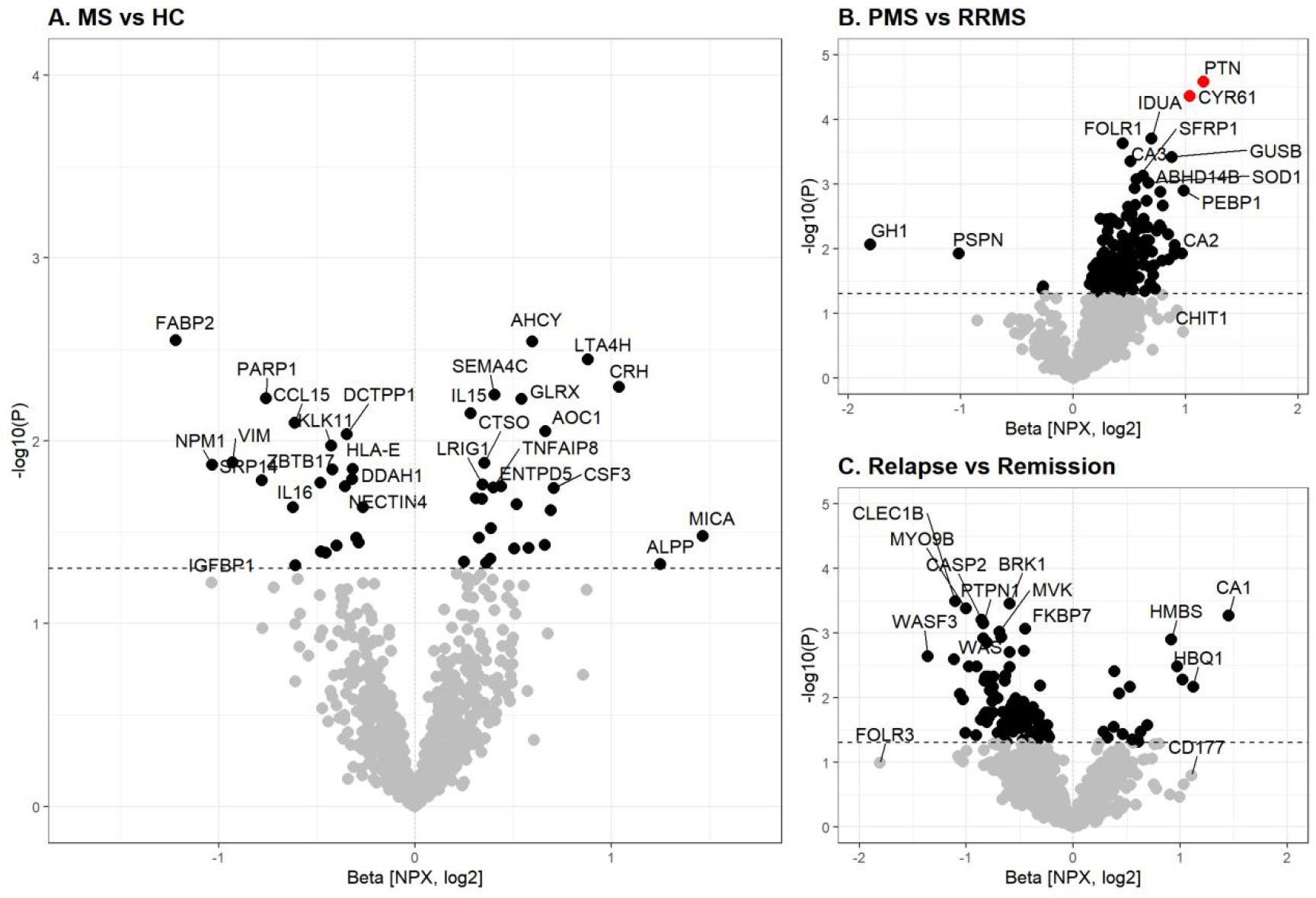
Plasma protein associations with multiple sclerosis and disease activity. Volcano plots illustrating plasma protein associations with [A] multiple sclerosis (MS) versus healthy controls (HC), [B] progressive (PMS) versus relapsing-remitting (RRMS) disease, and [C] relapse compared to remission at the time of sampling. Significance (P) and change in normalized protein expression (NPX) levels were determined using a multivariable linear regression model adjusted for sex, age at sampling, and sample handling. Significance of P<0.05 (black, dotted line) and P_FDR_<0.05 (red) are highlighted. Summary statistics are provided in **Supplementary Table 2a-c**.

High levels of CRNN and CXCL13 were associated with more severe disability at sampling (P<0.0001, P_FDR_<0.05), independent of progressive disease (**Figure 2**). CRNN was also suggestively associated with disease progression rate (P_MS_=0.16, P_RRMS_=0.059) and duration (P_MS_=0.036, P_RRMS_=0.056). LILRB5 levels were lower among those with more severe disease progression, although not significant after multiple test corrections. Low PTN and FOLR2 levels were associated with greater severity, particularly among RRMS (P=0.005). CTSF was positively correlated with disease duration (P=4.1×10^−5^, P_FDR_=0.044) but is likely due to secondary progressive disease (β=0.82, P=0.0030), as no correlation was observed among RRMS only (P=0.75). CRTAM was associated with EDSS (β=0.52, P=0.0087) and MSSS (β=0.48, P=0.0053) but not disease duration. In general, measures of disability and severity, including CRNN, CXCL13, and CTSF, did not differ between MS cases and controls.

**Figure 2.**
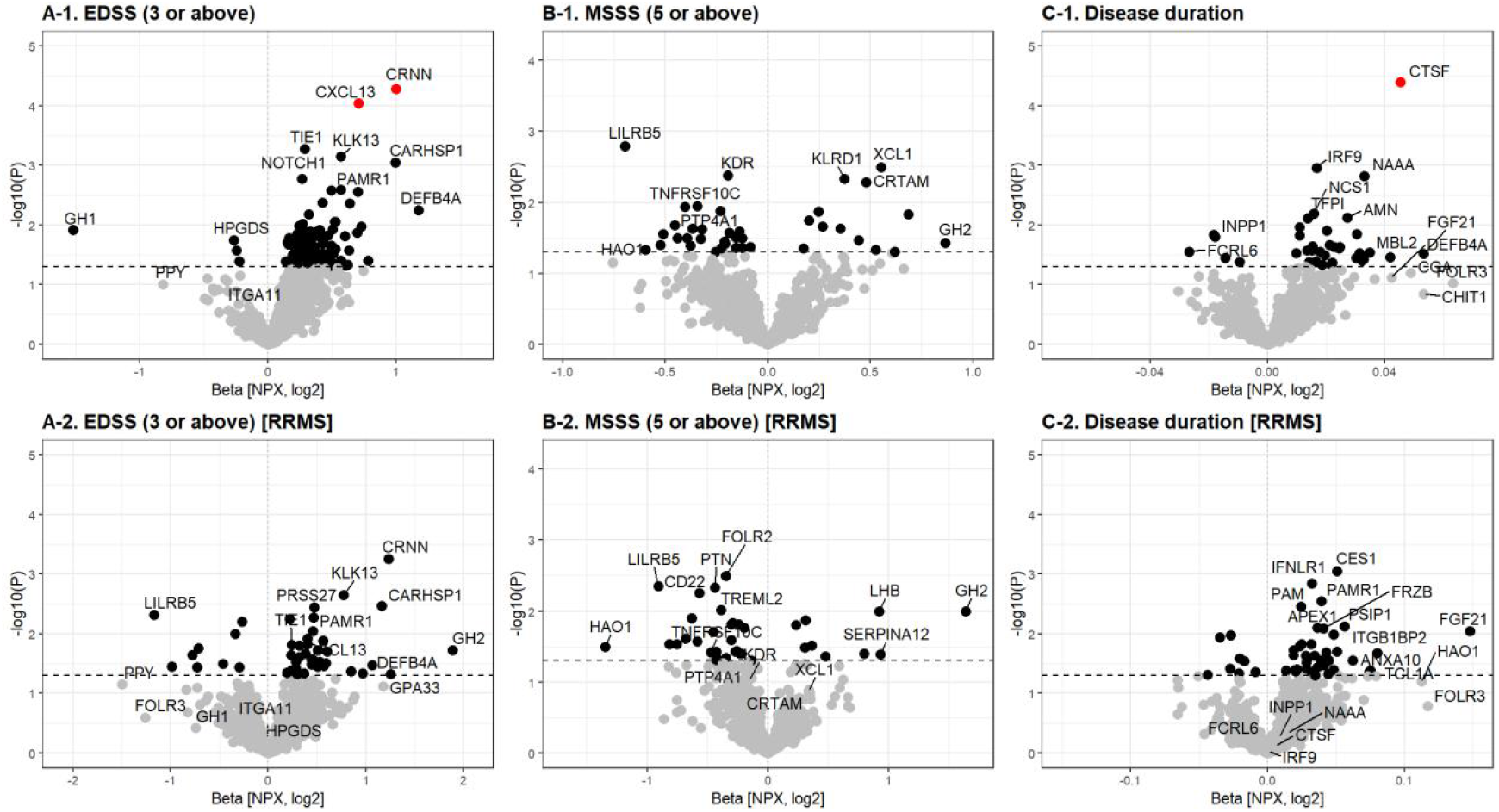
Plasma protein associations with multiple sclerosis disability and severity scores. Volcano plots illustrating plasma protein associations with [A] EDSS (≥ 3), [B] MSSS (≥ 5), and [C] disease duration (years) among all MS cases or only relapsing-remitting (RRMS) cases. Significance (P) and change in normalized protein expression (NPX) levels were determined using a multivariable linear regression model adjusted for sex, age at sampling, and sample handling. Significance of P<0.05 (black, dotted line) and P_FDR_<0.05 (red) are highlighted.

IgG index was associated with RRM2B levels in plasma, particularly among relapsing-remitting disease (β=0.47, P=1.7×10^−5^, P_FDR_=0.018; **S-Figure 2**). Similarly, CYR61 and FABP2 were positively correlated with IgG index (β=0.94, P=0.0012) despite FABP2 being lower in MS. No significant associations were observed with CSF mononuclear cell count or the number of MRI T2 lesions. However, PTN was suggestively associated with a higher number of MRI T2 lesions (β=0.68, P=0.012), likely due to association with progressive disease.

Although plasma NEFL was slightly higher in MS and associated with more MRI T2 lesion count (P=0.048), it was not significantly associated with MS, severity, or progressive course. Plasma NEFL was correlated with measures in CSF (r=0.76, P=1.3×10^−10^; **S-Figure 3**). TFF2 levels were positively correlated with NEFL in plasma (P=7.8×10^−5^, P_FDR_=0.043) and, to a lesser extent, CSF (P=0.12) (**S-Figure 4**).

## Discussion

We here conduct an exploratory profiling of the plasma proteome of MS cases using proximity extension technology, identifying several candidate biomarkers. One of which is a corticotropin-releasing hormone (CRH), which regulates stress response within the hypothalamic-pituitary-adrenal (HPA) axis. Although primarily expressed in the paraventricular nucleus of the hypothalamus, CRH controls peripheral inflammation through corticosteroid upregulation or directly by local CRH-receptor mediated stimulation of proinflammatory cytokines and lymphocyte proliferation^11^. The HPA axis also modulates the susceptibility and course of experimental autoimmune encephalomyelitis (EAE)^12^, the primary animal model for MS. In MS cases, increased CRH expression associated with hyperactivation of CRH/ vasopressin-expressing neurons has been observed postmortem and is impaired by active hypothalamic lesions corresponding to increased disease severity^13^. Immunosuppression of acute MS relapses by corticosteroids and the reduced inflammatory state typically associated with HPA-modulating conditions^11^, such as during pregnancy and intense exercise, further supports the involvement of CRH/HPA-axis in MS. Other disease-associated measures likely assess more systemic effects, including adenosylhomocysteinase (AHCY), an enzyme regulating transmethylation reactions associated with immune cell proliferation^14^, and intestinal/fatty acid-binding protein 2 (FABP2), a measure of intestinal permeability and epithelial damage^15^. However, their direct involvement in MS pathology and comorbidities remains unclear.

The primary associations were observed with progressive disease, particularly with pleiotrophin (PTN) and cysteine-rich angiogenic inducer 61 (CYR61/CCN1). PTN is a secreted neurotrophic factor and essential in the early differentiation of glial and neuronal progenitor cells. Although PTN expression is limited in adults, localized upregulation in the brain can occur after injury^16^, likely acting as a neuroprotective and regenerative factor^17^. PTN promotes neuronal survival and suppresses CNS inflammation in EAE as *Ptn* knockdown in astrocytes hinders recovery while late-phase intranasal administration of PTN improves disability^18^. This is further supported by the induction of PTN expression in demyelinated tissue, which promotes oligodendrocyte differentiation during remyelination^19^. These findings indicate PTN may not only be a predictive measure but a potential therapeutic target for MS, particularly progressive disease.

Similarly, CYR61 is also a secreted signaling protein associated with inflammation and tissue repair^20^. Although its involvement in MS is less clear, its regulatory function in the extracellular matrix during localized inflammation may affect fibronectin aggregation^21^, which inhibits remyelination efforts accelerating disease progression. Both PTN and CYR61 may be further complemented by measures of disability, including CXC chemokine 13 (CXCL13), a B-lymphocyte trafficking cytokine and established MS biomarker in CSF^2,22^; and cornulin (CRNN), a mediator of epidermal differentiation induced by proinflammatory cytokines^23^. However, cornulin is associated with Swedish snuff use^24^ and likely captures smoking history, causing an indirect association with disability.

This study benefits from proximity extension technology’s higher sensitivity and precision over conventional mass spectrometry or antibody-based methodologies^8^. However, this also increases susceptibility to bias such as from preanalytical variability^4,10^. In this study, we have performed additional corrections using standardized preprocessing markers and sensitivity analyses by sampling period but found no significant difference in the main findings. The cross-sectional study design limits temporal characterization, as differential levels could be either a risk or consequence of disease outcomes. However, limited association to disease duration would suggest early differentiation, an important factor in clinical applications.

In conclusion, we present a set of candidate biomarkers for MS, particularly progressive disease, which may be clinically relevant for characterizing disease and predicting response to immunomodulatory treatment. However, further investigation will be required to understand its mechanism in MS pathogenesis along with its predictive value for early detection and long-term monitoring of disease in a clinical setting.

**Table 1.**
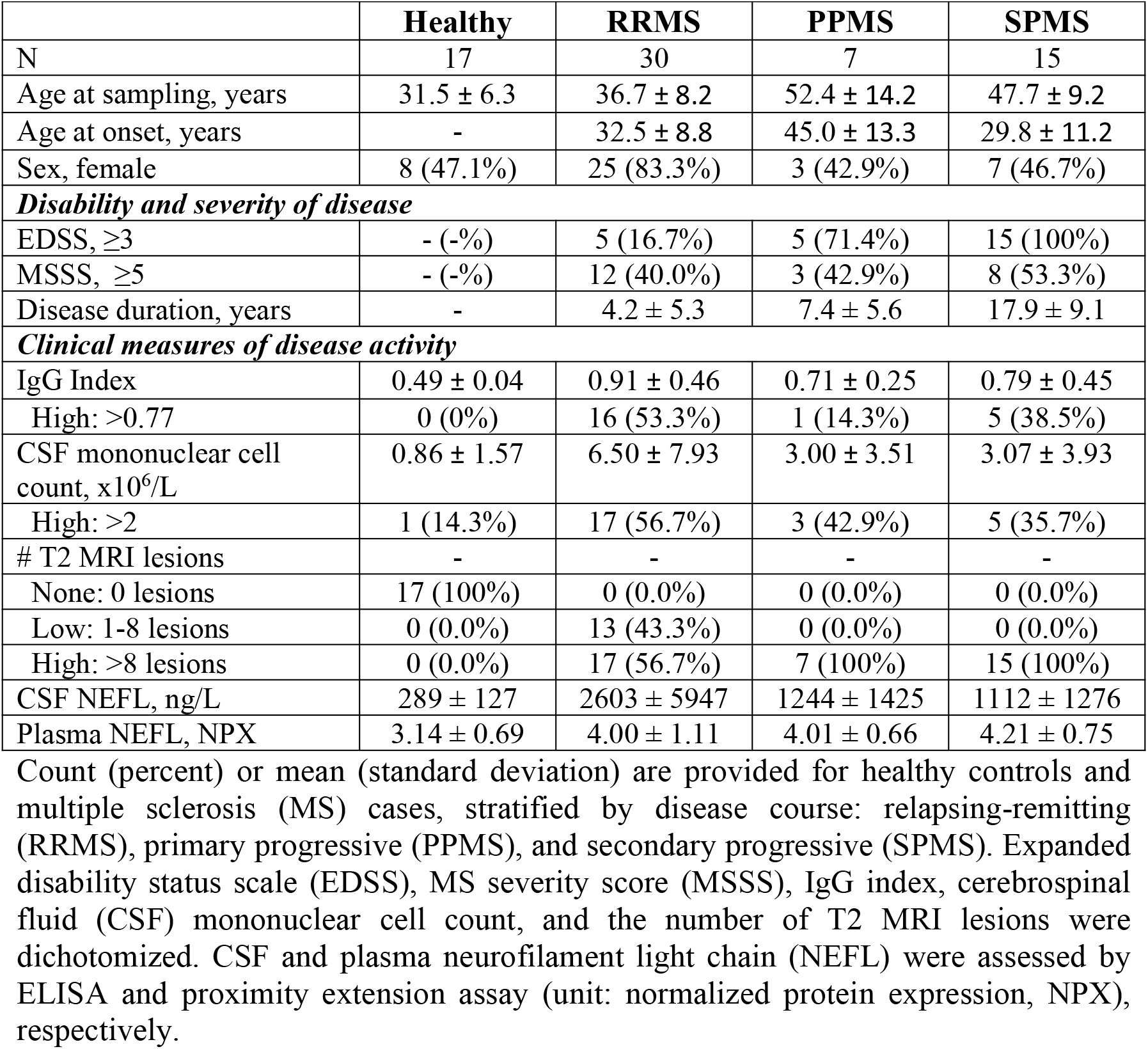
Descriptive statistics of cohort.

|  | **Healthy** | **RRMS** | **PPMS** | **SPMS** |
| --- | --- | --- | --- | --- |
| N | 17 | 30 | 7 | 15 |
| Age at sampling, years | 31.5 ± 6.3 | 36.7 ± 8.2 | 52.4 ± 14.2 | 47.7 ± 9.2 |
| Age at onset, years | - | 32.5 ± 8.8 | 45.0 ± 13.3 | 29.8 ± 11.2 |
| Sex, female | 8 (47.1%) | 25 (83.3%) | 3 (42.9%) | 7 (46.7%) |
| ***Disability and severity of disease*** |  |  |  |  |
| EDSS, ≥3 | - (-%) | 5 (16.7%) | 5 (71.4%) | 15 (100%) |
| MSSS, ≥5 | - (-%) | 12 (40.0%) | 3 (42.9%) | 8 (53.3%) |
| Disease duration, years | - | 4.2 ± 5.3 | 7.4 ± 5.6 | 17.9 ± 9.1 |
| ***Clinical measures of disease activity*** |  |  |  |  |
| IgG Index | 0.49 ± 0.04 | 0.91 ± 0.46 | 0.71 ± 0.25 | 0.79 ± 0.45 |
| High: >0.77 | 0 (0%) | 16 (53.3%) | 1 (14.3%) | 5 (38.5%) |
| CSF mononuclear cell count, x10<sup>6</sup>/L | 0.86 ± 1.57 | 6.50 ± 7.93 | 3.00 ± 3.51 | 3.07 ± 3.93 |
| High: >2 | 1 (14.3%) | 17 (56.7%) | 3 (42.9%) | 5 (35.7%) |
| # T2 MRI lesions | - | - | - | - |
| None: 0 lesions | 17 (100%) | 0 (0.0%) | 0 (0.0%) | 0 (0.0%) |
| Low: 1-8 lesions | 0 (0.0%) | 13 (43.3%) | 0 (0.0%) | 0 (0.0%) |
| High: >8 lesions | 0 (0.0%) | 17 (56.7%) | 7 (100%) | 15 (100%) |
| CSF NEFL, ng/L | 289 ± 127 | 2603 ± 5947 | 1244 ± 1425 | 1112 ± 1276 |
| Plasma NEFL, NPX | 3.14 ± 0.69 | 4.00 ± 1.11 | 4.01 ± 0.66 | 4.21 ± 0.75 |

Count (percent) or mean (standard deviation) are provided for healthy controls and multiple sclerosis (MS) cases, stratified by disease course: relapsing-remitting (RRMS), primary progressive (PPMS), and secondary progressive (SPMS). Expanded disability status scale (EDSS), MS severity score (MSSS), IgG index, cerebrospinal fluid (CSF) mononuclear cell count, and the number of T2 MRI lesions were dichotomized. CSF and plasma neurofilament light chain (NEFL) were assessed by ELISA and proximity extension assay (unit: normalized protein expression, NPX), respectively.

## Data Availability

Primary associations are provided in the supplements with all data provided in doi:10.17605/OSF.IO/9QT5H. Raw data and analytical scripts can be made available given reasonable request.

https://www.doi.org/10.17605/OSF.IO/9QT5H

## Abbreviations

AHCY: adenosylhomocysteinase
CRH: Corticotropin-releasing hormone
CRNN: Cornulin
CSF: Cerebrospinal fluid
CXCL13: Chemokine (C-X-C) ligand 13
CYR61/CCN1: Cysteine-rich angiogenic inducer 61
EAE: Experimental autoimmune encephalomyelitis
EDSS: Expanded disability status scale
FABP2: Fatty acid-binding protein 2
FDR: False discovery rate
HC: Healthy controls
HPA: Hypothalamic-pituitary-adrenal
MS: Multiple sclerosis
MSSS: Multiple sclerosis severity score
NEFL: Neurofilament light chain
NPX: Normalized protein expression
PEA: Proximity extension assay
PMS: Progressive multiple sclerosis
PPMS: Primary progressive multiple sclerosis
PTN: Pleiotrophin
RRMS: Relapsing-remitting multiple sclerosis
SPMS: Secondary progressive multiple sclerosis

## Acknowledgments

Protein quantification using proximity extension assays were performed at Olink Uppsala. This work was supported by the Swedish Research Council (Grant VR 2020-01638, 2020-02700), The Swedish Brain foundation, the Swedish Multiple Sclerosis Research Foundation, an endMS Doctoral Studentship (EGID:3045) from the Multiple Sclerosis Society of Canada and NEURO Sweden. J.H. and I.K. were partially supported by Horizon 2020 MultipleMS Grant 733161.

## Conflicts of interest

JH and IK have nothing to declare. FP has received research grants from Janssen, Merck KGaA and UCB, and fees for serving on DMC in clinical trials with Chugai, Lundbeck and Roche, and fees for expert witness statement for Novartis. TO has received lecture and/or advisory board honoraria, and unrestricted MS research grants from Astrazeneca, Biogen, Novartis, Merck, Roche, Almirall and Genzyme.

## Notes

### Author Declarations

The study was approved by the Stockholm Regional Ethical Review Board (no. 2009/2107-31/2, 2015/1280-32), and all participants have provided informed and written consent.

