## Supplementary Material for "Plasma protein profiling of multiple sclerosis using proximity extension assays"

**Supplementary Figure 1. Cross-validating plasma protein associations between discovery and replication partitions.**

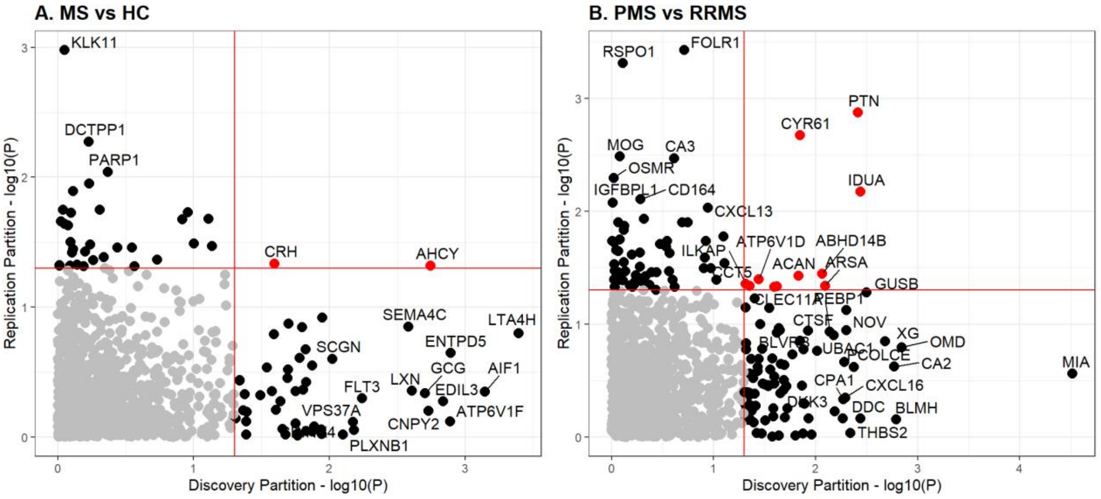


Significance (P) of plasma protein associations with [A] all multiple sclerosis (MS) cases versus healthy controls (HC) and [B] progressive MS (PMS) versus relapsing-remitting MS (RRMS), determined using a multivariable linear regression model adjusted for sex, age at sampling, and sample handling. Associations with $P<0.05$ are highlighted black (red line), while those with both $P<0.05$ (discovery/replication) are highlighted red. Discovery and replication partitions were determined with an equally distributed randomized selection by case/control or progressive/relapse-remitting status. These partitions were then used to conduct cross-validation and to identify primary features. Those with $P_{discovery}<0.05$ and $P_{replication}<0.05$ were selected, which include: [MSvsHC] CRH and AHCY; [PMSvsRRMS] PTN, CYR61, IDUA, ABHD14B, ARSA, ACAN, ATG4A, SFRP1, ATP6V1D, CCT5, and ILKAP.

**Supplementary Figure 2. Plasma protein associations with IgG index, cerebrospinal fluid (CSF) mononuclear cell count, and MRI T2 lesions.**

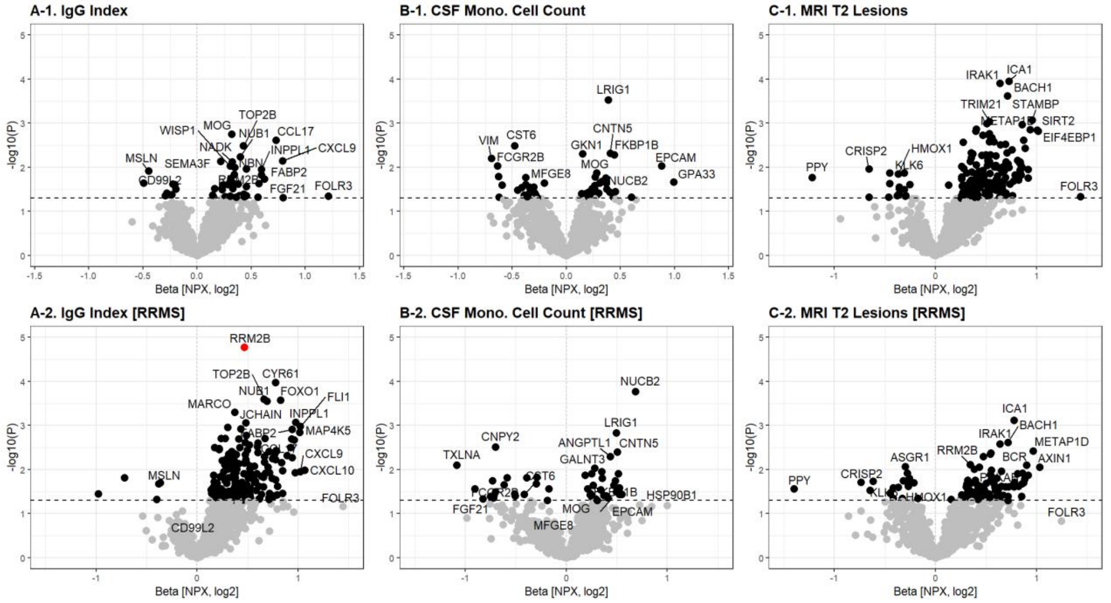


Volcano plots illustrating plasma protein associations with [A] IgG index (>0.77), [B] CSF mononuclear cell count (>2x10$^{6}$/L), [C] the number of T2 MRI lesions (>8) among all multiple sclerosis (MS) cases or only those with relapsing-remitting MS (RRMS). Significance (P) and change in normalized protein expression (NPX) levels were determined using a multivariable linear regression model adjusted for sex, age at sampling, and sample handling. Significance of P<0.05 (black, dotted line) and $P_{FDR}$<0.05 (red) are highlighted.

**Supplementary Figure 3. Correlation between cerebrospinal fluid (CSF) and plasma neurofilament light chain (NEFL) levels.**

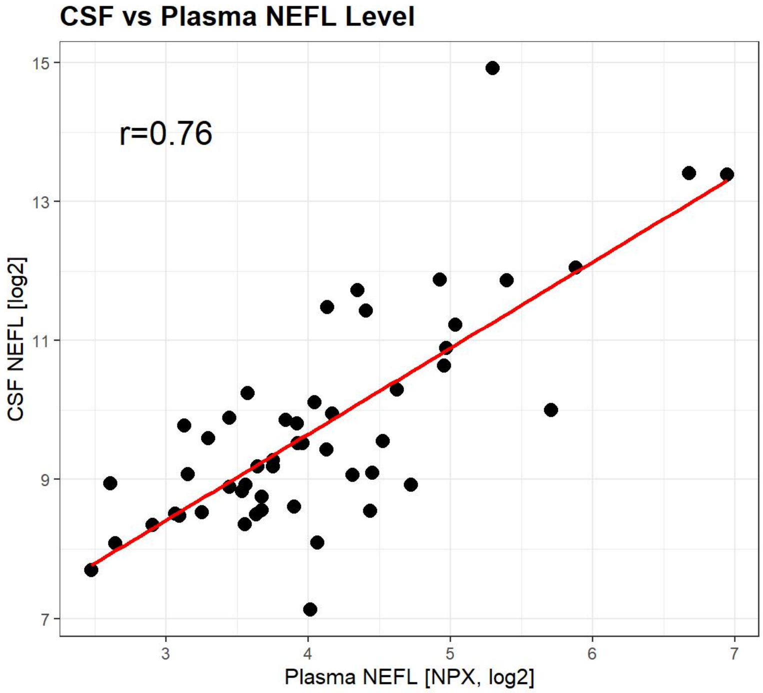


Scatterplot of CSF NEFL (ELISA, ng/L) and plasma NEFL (PEA, NPX) levels provided with line of best fit (red line) and Pearson's correlation coefficient (r).

**Supplementary Figure 4. Plasma protein associations with plasma and cerebrospinal fluid (CSF) neurofilament light chain (NEFL).**

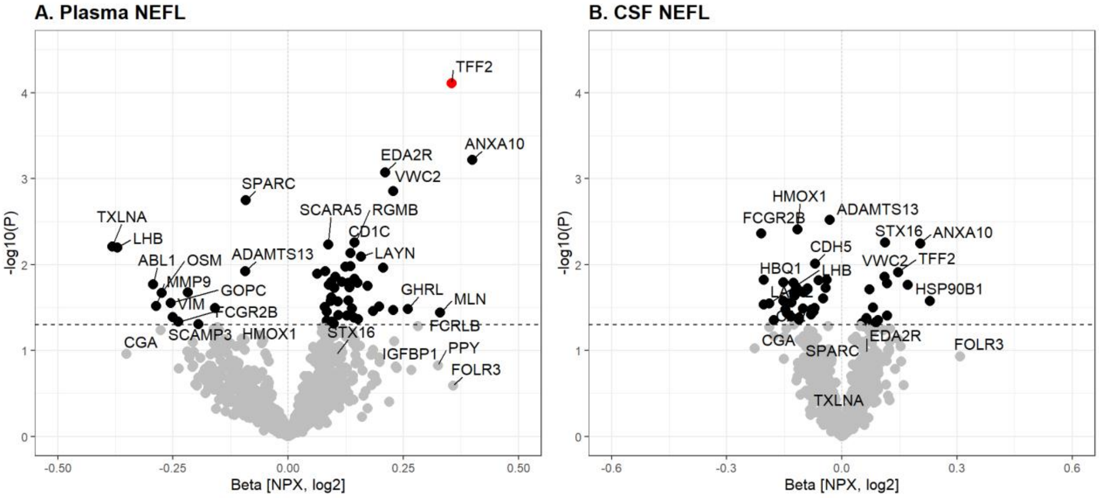


Volcano plots illustrating plasma protein association with [A] plasma and [B] CSF NEFL among all MS cases. Significance (P) and change in normalized protein expression (NPX) levels were determined using a multivariable linear regression model adjusted for sex, age at sampling, and sample handling. Significance of $P<0.05$ (black, dotted line) and $P_{FDR}<0.05$ (red) are highlighted.
